# Serum neopterin levels in relation to mild and severe COVID-19

**DOI:** 10.1101/2020.08.19.20178178

**Authors:** Josefina Robertson, Johanna M Gostner, Staffan Nilsson, Lars-Magnus Andersson, Dietmar Fuchs, Magnus Gisslen

**Author notes:** **Corresponding author:** Josefina Robertson, Department of Infectious Diseases, Sahlgrenska University Hospital, SE-416 85, Gothenburg, Sweden.

## Abstract

**Background:** The COVID-19 pandemic, caused by the coronavirus SARS-CoV-2, is rapidly spreading worldwide. There is limited information about prognostic markers that could help clinicians to identify COVID-19 patients with a poor prognosis. Serum levels of the immune activation marker neopterin has shown to be of prognostic value in patients with SARS. The aim of this study was to investigate whether serum neopterin is associated with the severity of COVID-19.

**Methods:** We included 34 patients with confirmed COVID-19 between March 3 and March 30, 2020. Fifteen patients had mild disease and did not require hospitalization, whereas 19 patients developed severe COVID-19 requiring intensive care. Concentrations of serum neopterin, tryptophan, and kynurenine were measured at and repeatedly after inclusion.

**Results:** We found a more than two-fold higher mean concentration of neopterin in severely ill patients (mean value 42.0 nmol/L (SD 18.2)) compared to patients with mild symptoms (16.9 nmol/L (SD 11.0)). All of the severe cases had elevated neopterin concentrations (>9.1 nmol/L) at the initial sampling with values ranging from 17.2 to 86.7 nmol/L. In comparison, 10 of 15 patients with mild disease had neopterin levels above 9.1 nmol/L, with concentrations in the range from 4.9 to 31.6 nmol/L. Neopterin levels gradually decreased during the course of COVID-19, but severe cases maintained elevated levels for a longer period. Moreover, lower levels of tryptophan and higher levels of kynurenine, indicating an increased tryptophan catabolism, were seen in the group with severe cases.

**Conclusions:** In conclusion, we found that serum neopterin levels are associated with the severity of COVID-19. Our findings suggest that neopterin could be used as a prognostic marker, but further studies are needed to elucidate how it can be used in clinical praxis.

## Introduction

The COVID-19 pandemic, caused by the coronavirus SARS-CoV-2, is rapidly spreading worldwide (1). Most patients have mild symptoms from the upper respiratory tract, whereas a minor, but not negligible proportion, suffers from a severe form of the disease which in some cases require intensive care (2). There is limited information about prognostic markers that could help clinicians to identify COVID-19 patients with a poor prognosis. During the outbreak of the severe acute respiratory syndrome (SARS) in 2002–2003, caused by the similar coronavirus SARS-CoV, levels of the immune activation marker neopterin were found to predict the course of disease (3).

Neopterin (6-(D-erythro-1′, 2′, 3′-trihydroxypropyl)-pterin) is a well-established immune activation marker with elevated concentrations seen in many inflammatory states including infections, autoimmune disorders, and cancer (4). In acute viral infections such as hepatitis (5), Cytomegalovirus disease (6), Rubella (7), and dengue fever (8), serum neopterin levels correlate with the activity of the disease, and can be detected before antibody production (4). The elevation of neopterin originates mainly from the increased synthesis by human monocyte-derived macrophages, whereby interferon-gamma (IFN-y) is the most central activating cytokine (9). IFN-y also promotes the conversion of the essential amino acid L-tryptophan (TRP) to N-formylkynurenine, which is rapidly converted into the more stable kynurenine (KYN), by induction of the anti-proliferative and immunoregulatory enzyme indoleamine 2,3-dioxygenase (IDO). The first step of the TRP breakdown is the rate-limiting step in the TRP catabolic route along the KYN axis, and the KYN to TRP ratio can be used as a measure of the IDO enzyme activity (10). Taken together, IFN-y mediated immune response to viral infections may lead to elevated neopterin levels, as well as increased TRP degradation and elevated KYN to TRP ratio (4, 11).

In patients with SARS, elevated levels of neopterin were detected already at the day of symptom onset, and rose to a maximum level at day 3 (3). Moreover, patients with higher levels of neopterin at an early stage suffered from a severer course of disease, including higher and longer period of fever, more severe dyspnea, longer hospitalization, and more complications (3). Considering these findings and the similarity of the coronaviruses, neopterin may be a useful prognostic marker for the course of COVID-19. The aim of this study was to investigate whether serum neopterin levels in COVID-19 patients are associated with the severity of disease.

## Methods

### Participants

We included 34 patients with COVID-19 who were admitted to the Department of Infectious Diseases at the Sahlgrenska University Hospital, Gothenburg, Sweden. All cases were confirmed with reverse transcriptase polymerase chain reaction (RT-PCR) from nasopharyngeal and throat aspirates. Fifteen patients had mild disease and did not require hospitalization, whereas 19 patients developed severe COVID-19 and required intensive care. Blood samples were collected between March 3 and March 30, 2020.

### Serum neopterin, TRP, and KYN measurements

Serum neopterin concentrations were determined using enzyme-linked immunosorbent assay (ELISA) (BRAHMS GmbH, Hennigsdorf, Germany) as described by the manufacturer’s instructions. Sensitivity of the test was 2 nmol/L neopterin. The upper normal reference level was 9.1 nmol/L in serum (12). Serum TRP and KYN concentrations were measured by a reverse-phase HPLC method (13), using a Varian ProStar HPLC system equipped with a solvent delivery module (model 210), an autosampler (model 400, both Varian ProStar), an UV-spectrometric detector (SPD-6A, Shimadzu), and a fluorescence detector (model 360, Varian ProStar). Varian Star Chromatography Workstation (version 6.30) software was used.

### Statistical analyses

Descriptive statistics are shown for all variables involved in the analyses, presented as means with standard deviations (Table 1). Continuous variables were log_10_ transformed. Student’s ttest was used for group comparisons. Changes in log concentrations from first to last measure were analyzed with paired t-test. Associations were measured with Pearson correlation. All statistical analyses were performed with the Statistical Package for the Social Sciences (SPSS) software version 25 (SPSS, Chicago, Illinois, USA) or Prism (GraphPad software version 8.0, La Jolla, California, USA). A significance level below 0.05 was considered as statistically significant.

**Table 1.**
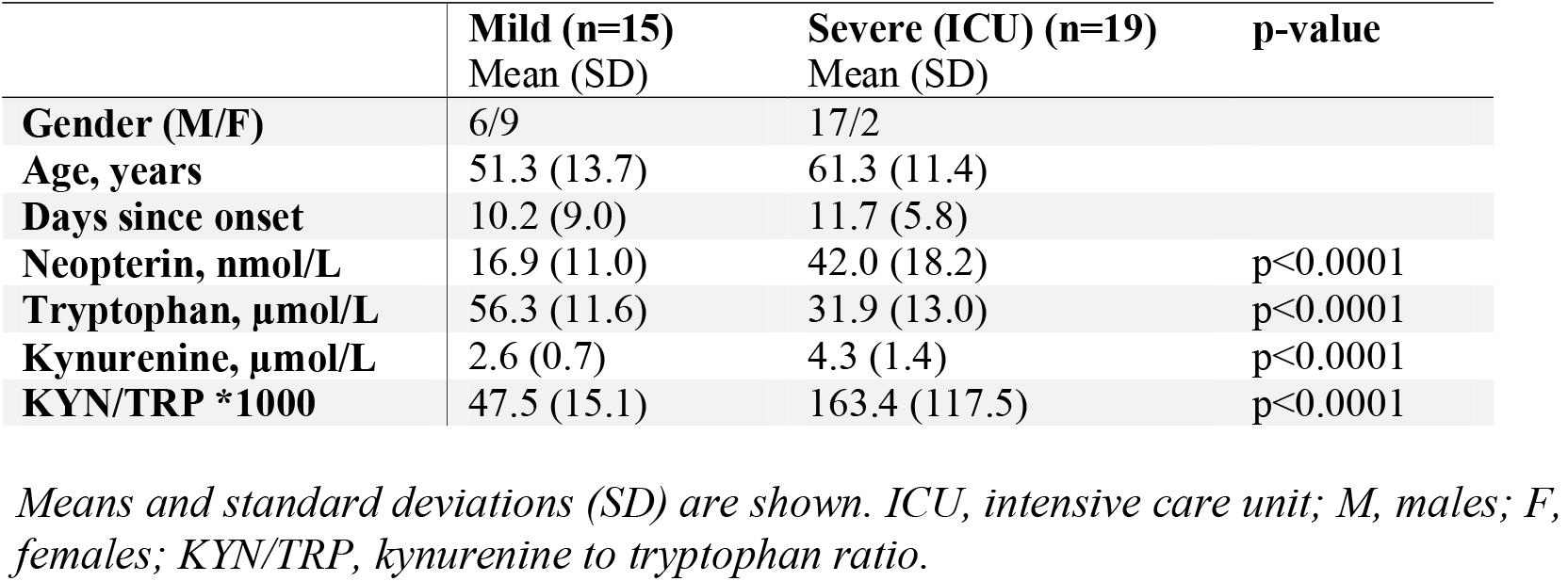
Demographic data and concentrations of neopterin and amino acids from the study population divided into two groups based on disease severity.

|  | **Mild (n=15)**<br>Mean (SD) | **Severe (ICU) (n=19)**<br>Mean (SD) | **p-value** |
| --- | --- | --- | --- |
| **Gender (M/F)** | 6/9 | 17/2 |  |
| **Age, years** | 51.3 (13.7) | 61.3 (11.4) |  |
| **Days since onset** | 10.2 (9.0) | 11.7 (5.8) |  |
| **Neopterin, nmol/L** | 16.9 (11.0) | 42.0 (18.2) | p<0.0001 |
| **Tryptophan, µmol/L** | 56.3 (11.6) | 31.9 (13.0) | p<0.0001 |
| **Kynurenine, µmol/L** | 2.6 (0.7) | 4.3 (1.4) | p<0.0001 |
| **KYN/TRP \*1000** | 47.5 (15.1) | 163.4 (117.5) | p<0.0001 |

*Means and standard deviations (SD) are shown. ICU, intensive care unit; M, males; F, females; KYN/TRP, kynurenine to tryptophan ratio.*

## Results

The study population comprised 34 patients, of which 15 displayed mild symptoms and 19 developed a severe form of COVID-19. The mean age among patients with mild symptoms was 51.3 years (SD 13.7), and 61.3 years (SD 11.4) in the group with severe symptoms.

To test whether neopterin levels are associated with the severity of COVID-19, we compared serum neopterin concentrations between patients with mild and severe disease. We found a more than two-fold higher mean concentration of neopterin in severely ill patients (mean value 42.0 nmol/L (SD 18.2)) compared to patients with mild symptoms (16.9 nmol/L (SD 11.0)) (Table 1, Figure 1 and 2). All of the severe cases had elevated neopterin concentrations (>9.1 nmol/L) at the initial sampling with values ranging from 17.2 to 86.7 nmol/L, measured at day 4–20 (mean 11.7 (SD 5.8); median 14) since onset of symptoms. In comparison, 10 of 15 patients with mild disease had neopterin levels above 9.1 nmol/L at day 2–19 (mean 10.2 (SD 9); median 9), with concentrations for the entire group in the range from 4.9 to 31.6 nmol/L. These results show that patients with severe COVID-19 display higher levels of neopterin than mild cases. Since renal insufficiency may affect serum neopterin levels (14), an analysis of the correlation between creatinine and neopterin levels in severe cases was performed. No significant correlation was found (r = 0.36, p = 0.13).

**Figure 1.**
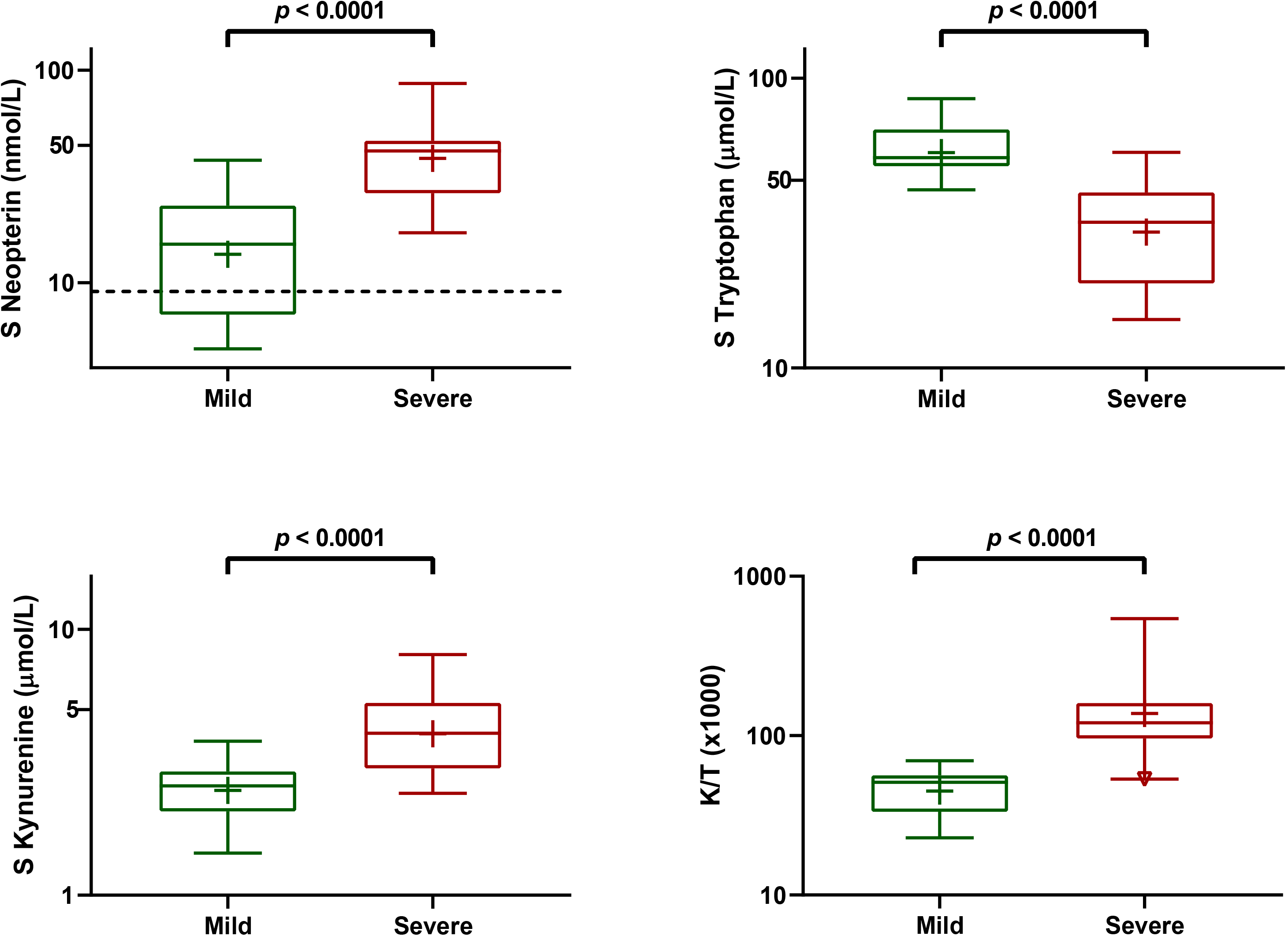
Concentrations of serum neopterin, tryptophan, and kynurenine, as well as kynurenine to tryptophan ratio (K/T) in patients with mild (green) and severe (red) form of COVID-19 (n=34). The dotted line represents the upper normal reference limit of neopterin at 9.1 nmol/L.

**Figure 2.**
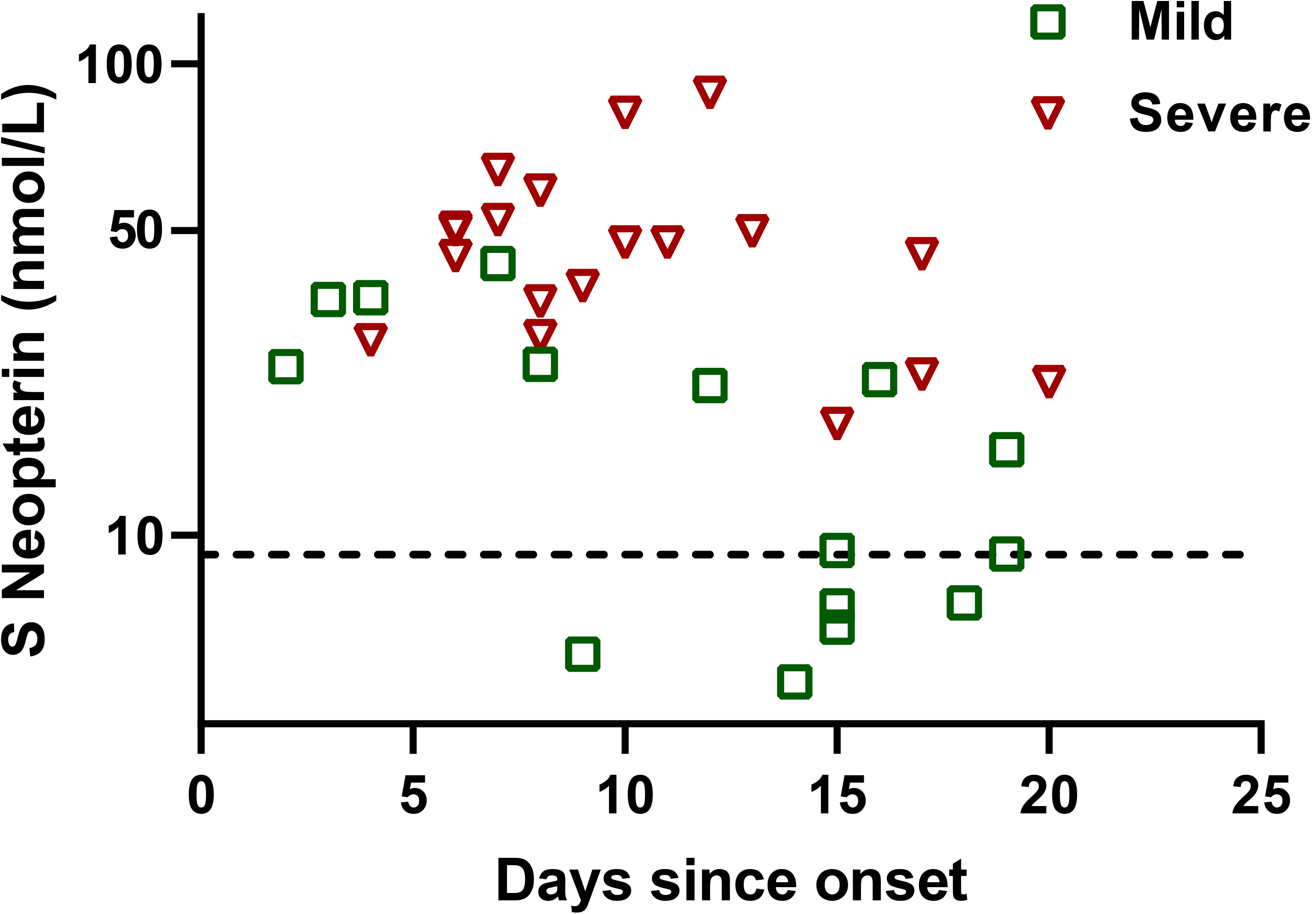
Serum neopterin concentrations in patients with mild (green) and severe (red) form of COVID-19 (n=34). The dotted line represents the upper normal reference limit of neopterin at 9.1 nmol/L.

As a next step, we studied the trajectory of neopterin levels during the progression of COVID-19 by repeated measurements. We found that serum neopterin levels decreased over time regardless of mild or severe disease (p < 0.0001, Figure 3). However, the group with severe disease displayed higher concentrations than the patients with mild disease throughout the study period (Figure 3). Twelve of the patients with mild disease had normal levels (<9.1 nmol/L) of neopterin at the last measurement. In contrast, only one patient among the severely ill returned to a normal level during the study period (Figure 3). These results show that neopterin levels gradually decrease during the course of COVID-19, but that severe cases maintain elevated levels for a longer period. Collectively, our findings indicate an association between neopterin and the severity of COVID-19.

**Figure 3.**
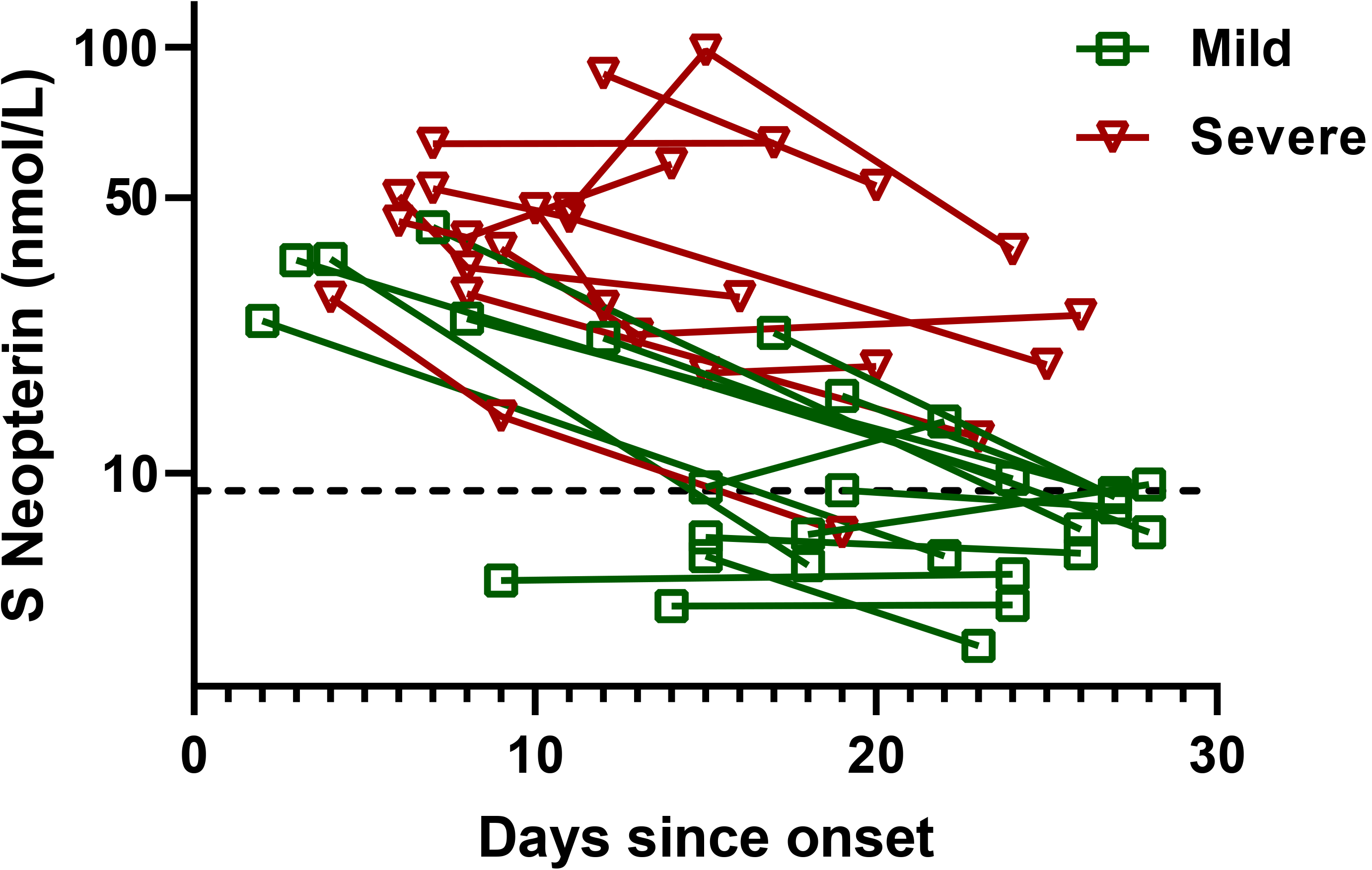
Repeated measurements of serum neopterin concentrations in patients with mild (green) and severe (red) form of COVID-19 (n=34). The dotted line represents the upper normal reference limit of neopterin at 9.1 nmol/L.

Furthermore, we investigated TRP and KYN concentrations in relation to mild and severe COVID-19. As seen in figure 1, we found lower levels of TRP and higher levels of KYN in severe cases, as compared to mild cases. This indicates that a severe form of COVID-19 is associated with an increased TRP catabolism. Additionally, elevated KYN/TRP concentrations closely correlated to neopterin concentration (r = 0.7, p < 0.0001) (Figure 4).

**Figure 4.**
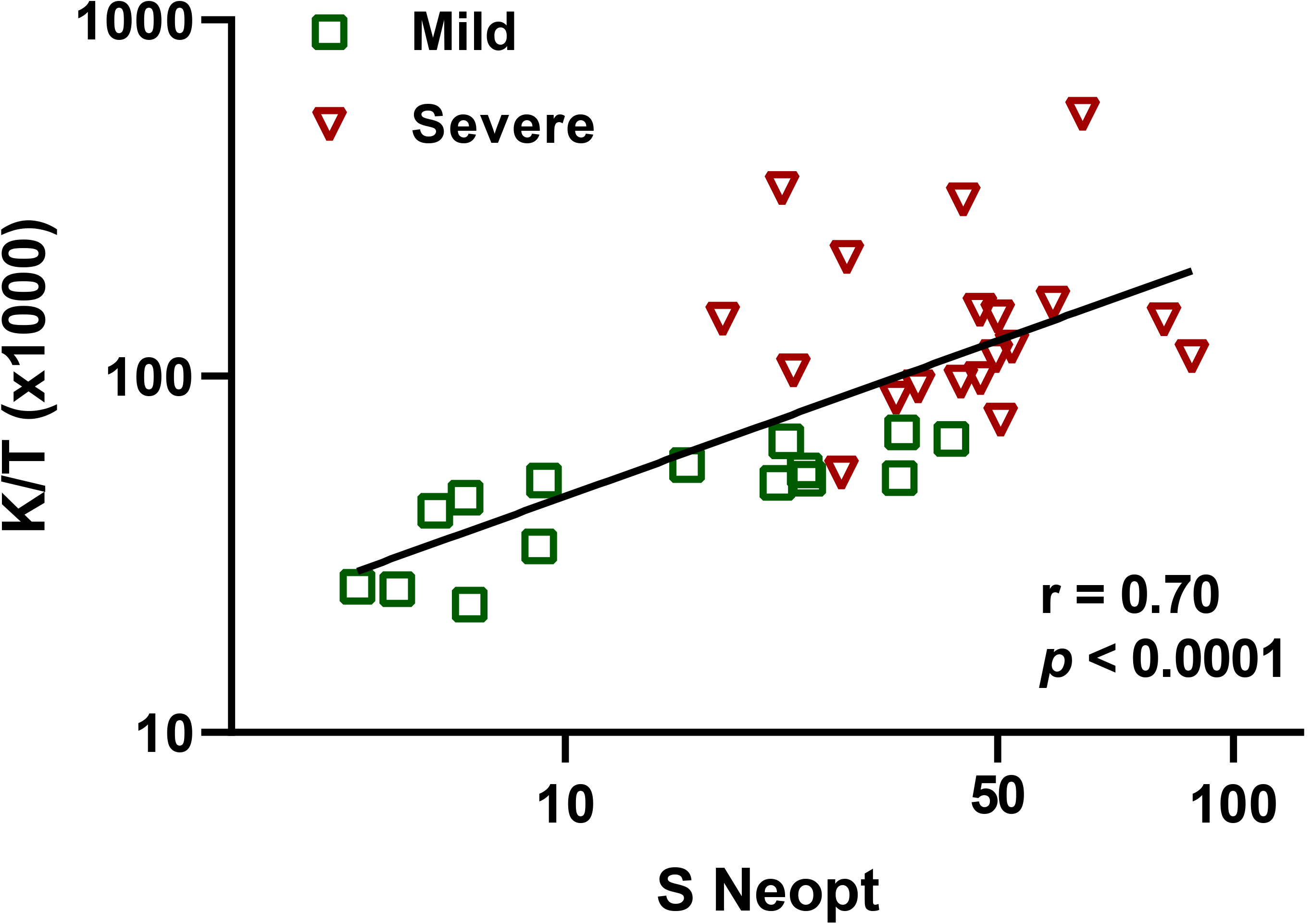
The correlation between serum neopterin (Neopt) concentration and kynurenine to tryptophan ratio (K/T) in patients suffering from COVID-19 (n=34).

## Discussion

In the present study, we investigated if serum neopterin levels in patients with COVID-19 are associated with the severity of disease. We report that higher neopterin concentrations were seen in patients who developed severe disease compared to patients with mild disease. In the group with severe COVID-19, we also found an increased metabolism of TRP, as expressed by higher KYN to TRP ratio.

The observed difference in average neopterin concentrations between mild and severe COVID-19 is in agreement with previous results from SARS patients (3). In that study, higher neopterin levels were associated with a longer fever period, as well as a severer course of disease. It is also in line with prior research on other viral infections (5, 8, 15). The elevated neopterin levels in severely ill patients with COVID-19 indicate a potent activation of monocytes/macrophages during the symptomatic period (9). Moreover, our detection of high neopterin concentrations already at day 2 after symptom onset suggests an early inflammatory response to SARS-CoV-2. Likewise, the study of SARS patients (3), and also a study of dengue fever (8), recognized neopterin elevation at the first day of symptoms. The characteristics of neopterin being elevated at an early stage of disease, as well as being associated to disease severity, suggest it as a useful prognostic marker for COVID-19.

In the present study, most patients with mild symptoms returned to normal neopterin levels at the end of the study period. In contrast, neopterin levels remained elevated during the course of disease in the severely ill. This probably illustrates a more advanced and prolonged inflammatory state, as suggested by others (16).

The increased KYN to TRP ratio in patients with severe COVID-19 reflects an increased TRP catabolism. Similar findings have been presented for HIV-infected patients with a progressive disease (17). TRP deprivation is an effective strategy of the Th1-type immune response to reduce undesirable proliferation of pathogens and infected cells (18), which may be useful for disease control in COVID-19.

When interpreting our results, one must consider the potential involvement of renal function (14). Acute kidney injury has been found in 50% of fatal COVID-19 cases (19). In this context, the lack of creatinine measurements in mild cases make out a limitation of the present study. However, we found no correlation between creatinine and neopterin levels in the severe cases at the first measurement.

## Conclusions

Our study shows an association between neopterin levels and severity of COVID-19, and also elevated concentrations early in disease progression. In conclusion, serum neopterin is a potential marker for the prognosis of COVID-19 when detected in blood samples from a few days since symptom onset, but further studies are needed to elucidate how it can be used in clinical praxis.

## Data Availability

The datasets used and/or analyzed during the current study are available from the corresponding author on reasonable request.

## List of abbreviations

SARS: severe acute respiratory syndrome
IFN-y: interferon-gamma
TRP: tryptophan
KYN: kynurenine
IDO: indoleamine 2,3-dioxygenase
RT-PCR: reverse transcriptase polymerase chain reaction
ELISA: enzyme-linked immunosorbent assay

## Declarations

### Ethics approval and consent to participate

The study has been approved by the Swedish Ethical Review Authority (2020–01771). Participants were enrolled after informed consent.

### Consent for publication

Not applicable.

### Authors’ contributions

JR and MG were responsible for the conception and design of the study, as well as for acquisition and analysis of data. JMG and DF performed the biochemical analyses. MG, JR, and SN performed the statistical analyses. All authors took part in drafting the manuscript and approved the final version.

